# Modeling the impact of prioritizing first or second vaccine doses during the 2022 mpox outbreak

**DOI:** 10.1101/2023.10.13.23297005

**Authors:** Patrick A. Clay, Emily D. Pollock, Enrique M. Saldarriaga, Preeti Pathela, Michelle Macaraig, Jane R. Zucker, Bindy Crouch, Ian Kracalik, Sevgi O. Aral, Ian H. Spicknall

## Abstract

**Background:** Early in the 2022 mpox outbreak, vaccine doses and administrative capacity were limited. The US recommendation was to administer two doses of the JYNNEOS® vaccine 4 weeks apart. However, because of the limited vaccine supply and high demand, New York City (NYC) prioritized administration of first doses to reach a larger portion of the impacted population as quickly as possible. We estimated mpox cases averted compared to strategies that prioritized 2-dose vaccination for a smaller portion of the population.

**Methods:** We fit a dynamic network transmission model to incident mpox cases reported by NYC, as well as to first and second vaccine doses administered from May 2022 through March 2023. Model output consisted of predicted cases over time when vaccine doses were administered with the ‘first-dose priority’ strategy, compared with counterfactual simulations where individuals were either pre-allocated full courses of the vaccine (‘second-dose priority’ strategy), or not pre-allocated doses, but where doses were administered to those eligible for a second dose ahead of those waiting for a first dose (‘intermediate’ strategy).

**Results:** We estimate that NYC’s ‘first-dose priority’ strategy averted 81% [IQR:75%–86] of potential mpox cases. Their ‘first-dose priority’ strategy was more effective than alternatives, averting 3.0% [IQR:1.2%–4.5%] more cases than the ‘intermediate’ strategy, and 9.5% [IQR:7.7%–12%] more cases than the ‘second-dose priority’ strategy.

**Conclusions:** A focus on widespread, 1 dose vaccination during future mpox outbreaks can reduce cases and limit transmission in scenarios of limited vaccine supply, limited vaccine administration capacity, or increased demand.

## Introduction

Over 30,000 individuals in the United States were diagnosed with mpox in 2022, with 33% of these cases occurring in May through July, and daily cases peaking on August 1^st^. Official CDC guidance was to administer two doses of the JYNNEOS® vaccine 4 weeks apart to priority populations (primarily gay, bisexual, and other men who have sex with men, hereafter referred to as MSM, with multiple recent sexual partners) [1]. During this period of rapidly increasing daily cases, demand for vaccination exceeded available doses of the JYNNEOS® vaccine and the capacity for administration in many jurisdictions. Given the rapidly increasing number of cases in New York City (NYC) and the resultant need to protect priority populations as quickly as possible, NYC focused their vaccination campaign on widespread, 1 dose vaccination, (a ‘first-dose priority’ strategy), with plans to administer second doses when supply and demand allowed, rather than following official CDC recommendations, which would have resulted in earlier 2 dose vaccination in a narrower portion of the population. Due to the urgency of the outbreak, NYC Health was required to make this decision with limited data on the effectiveness of 2 dose vs. 1 dose vaccination.

## Methods

We used an SEIR (susceptible, exposed, infectious, resistant) natural history in our model, expanded to account for pre-symptomatic transmission and multi-dose vaccination [6]. Individuals start in the susceptible (*S*) class. An infectious individual has a probability of infecting a susceptible partner given by the probability of sexual contact per timestep multiplied by the probability of infection per sexual exposure (*μ*). Upon acquiring infection, individuals enter a pre-symptomatic non-infectious state (*E*), after which they have a probability per day of entering the pre-symptomatic and infectious state (*P*), then the symptomatic and infectious state (*I*), and finally the recovered and resistant state (*R*). Individuals can also be vaccinated with 1 (*V*1) or 2 (*V*2) doses of the vaccine, with effects described below. Symptom onset reduces sexual contact rate with ‘main’ and ‘casual’ partners by 50% and causes individuals to move to one lower sexual activity level until they recover. A proportion of individuals, (*ρ*), will test for mpox *γ* days after they develop symptoms, with *γ* based on the date-varying time between reported symptom onset and mpox tests in case data. After a positive mpox test, individuals become aware of their infection status, and no longer transmit the virus through sexual activity. Individuals who learn their infection status sustain contact with their ‘main’ partner such that they have a 10% chance of infecting their ‘main’ partner over the duration of the infection, reflecting prior estimates of household transmission [7]. Infected individuals who seek medical care are reported as diagnosed cases, hereafter referred to as *cases*.

We initiated the model by introducing 5 newly exposed individuals into the two highest sexual activity groups on May 14^th^. This produced a median of 5 cumulative cases in the model on May 31^st^, matching case data from NYC.

Our network does not account for potential periods of increased sexual activity associated with the high frequency of MSM gatherings (e.g., pride festivals) throughout the summer. Additionally, we assume that importation of mpox cases from countries impacted by mpox prior to the outbreak in the United States may have fed the initial surge of mpox cases. Thus, we model a ‘surge period’ during which we randomly select individuals in the top two sexual activity groups to enter the *E* state via ‘extra-network’ contacts at a rate fit to incident case report data (see fitting procedure below). We set this surge period to occur in the two weeks after NYC Pride (June 26^th^ – July 10^th^) but note that there were many MSM oriented gatherings in NYC at this time and are not assuming that a disproportionate level of exposures happened at any specific event.

Over the course of each simulation, individuals decrease their probability of one-time partnerships and their rate of sexual contact with casual partners as a function of perceived risk of mpox [8]. We parameterize this perceived risk based on frequency of mpox discussion on online LGBT+ discussion forums over time, as discussed by Clay et al. [4], and fit the magnitude of this behavioral adaptation to incident case report data (see fitting procedure below).

### Vaccination

We ran three vaccination scenarios (Figure 1). We first ran a scenario where first and second doses administered per week matched those reported by NYC through March 18, 2023. This scenario is our baseline ‘first-dose priority’ strategy, as prioritizing widespread, 1 dose vaccination was the goal of NYC’s vaccination campaign. We then ran a counterfactual scenario where the total doses administered per week matched those in our ‘first-dose priority’ strategy, but during each timestep, doses were always administered first to individuals who were due for a second dose, and first doses were only given once there were no individuals waiting for a second dose (‘intermediate’ strategy). Finally, we ran an additional counterfactual scenario where individuals receiving their first dose were pre-allocated a second dose to be administered five weeks later, reflecting average time between doses in other jurisdictions (appendix). Thus, every individual completed their course on time, but total dose administration was slowed (‘second-dose priority’ strategy). The cumulative number of cases averted by vaccine administration was calculated as the cumulative number of cases in the ‘no vaccination’ scenario minus the cumulative cases in a scenario with vaccine administration. We compared the number of cases averted by the baseline scenario to the number of cases averted by the counterfactual vaccination scenarios.

**Figure 1:**
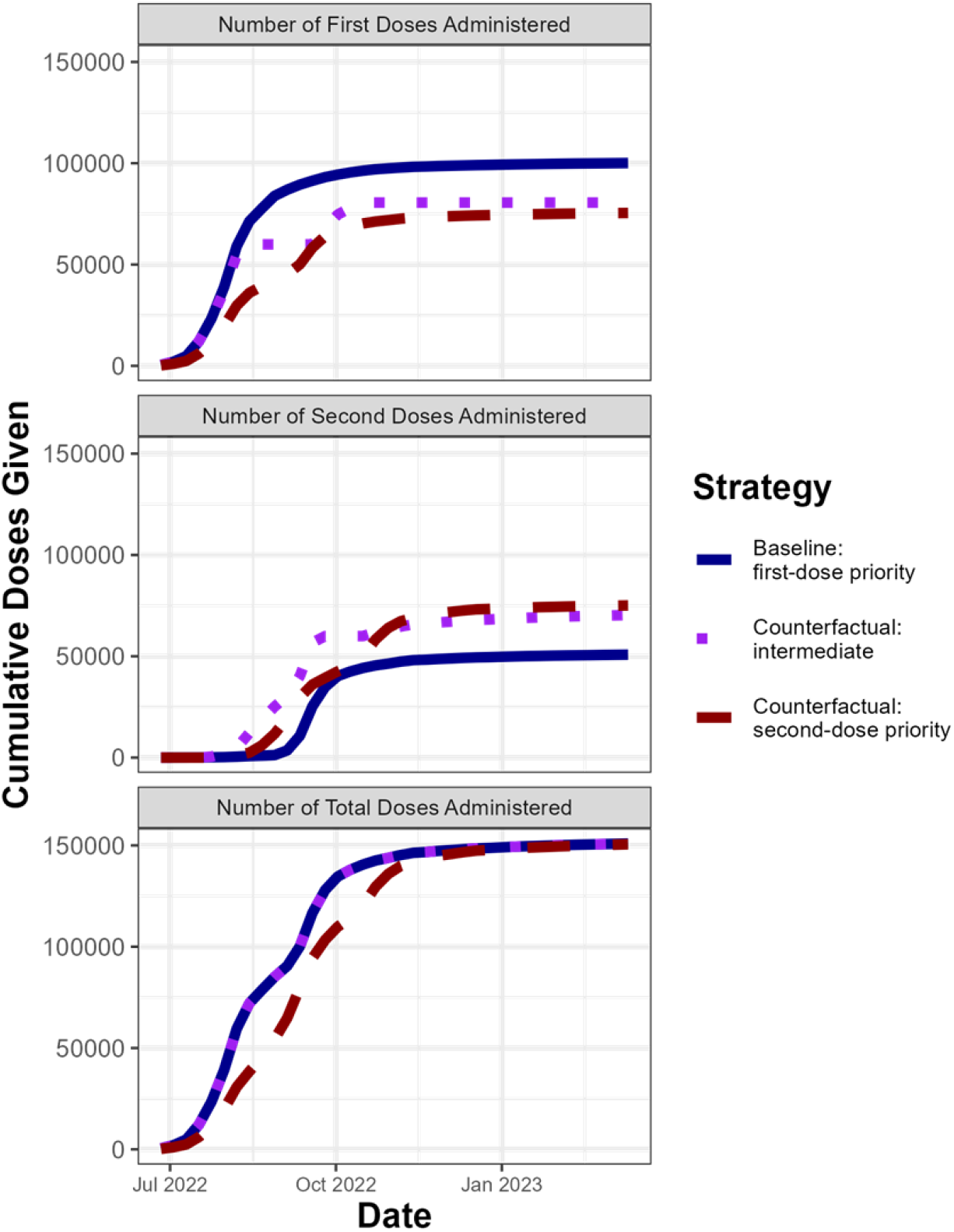
Allocation of doses over time for each vaccine administration strategy*. Y-axis shows number of doses given over time on the X-axis. Top two panels indicate number of first and second doses given, respectively, for each vaccine administration strategy while bottom panel indicates the sum of first and second doses given for each vaccine administration strategy. Colors indicate different vaccine administration strategies. *First-dose priority: first and second doses in model are based on first and second doses administered by NYC. Intermediate: Individuals who are eligible for a second dose receive priority for available doses, but there is no preallocation. Second-dose priority: doses are preallocated to ensure that all individuals receive full course of thes vaccine.

While MSM, transgender, gender non-conforming and non-binary adults (TGNCNB) with multiple sexual partners were all initially eligible for mpox vaccination, we only model MSM due to availability of sexual network data [5]. According to a 2020 citywide population-based survey, MSM made up 82% of MSM and TGNCNB adults in NYC with 2+ sexual partners over a 12-month period [9]. Thus, we assume that 82% of mpox vaccines administered in NYC were administered to MSM in our model.

As vaccines were originally intended for selected groups, including individuals with multiple recent sexual partners [10], we only vaccinate individuals in the top two sexual activity groups for the first four weeks of vaccination, only vaccinate individuals in the top four sexual activity groups for the next four weeks of vaccination, and only vaccinate individuals with a non-zero probability of engaging in one-time sexual partnerships for the rest of the simulation. In our model, susceptible (*S*) and pre-symptomatic (*E*, *P*) individuals are eligible for vaccination. However, vaccination does not prevent individuals in the *E* or *P* class from becoming infectious. Individuals are eligible for second doses of the vaccine if they have received their first dose at least four weeks in the past. In the case of breakthrough infections (i.e. infections of vaccinated individuals), we assume these individuals enter the pre-symptomatic (*E*) class, i.e., prior vaccination has no effects on subsequent contagiousness or pathogenicity.

CDC recently published JYNNEOS® effectiveness estimates (the overall reduction in infection risk for a vaccinated individual compared to an unvaccinated individual over an observed period) of 75.2% for one dose, and 85.9% for two doses [11]. We use these values in our model as per-exposure reductions in transmission probability due to vaccination, hereafter referred to as per-exposure vaccine efficacy. Setting vaccine effectiveness equal to per-exposure vaccine efficacy requires the assumption that individuals in the vaccine effectiveness study were not exposed to mpox multiple times (Figure S2). If individuals in the vaccine effectiveness study were exposed to mpox multiple times, we would expect for per-exposure vaccine efficacy to be greater than vaccine effectiveness. We thus repeated our analysis while increasing per-exposure vaccine efficacy by 10% and found no qualitative difference in our results (Figures S4-6). In all scenarios, we assume that doses become effective two weeks after administration.

### Fitting Process

Our model has three free parameters whose values were informed via fitting: (1) the probability of transmission per exposure (‘transmission probability’, *μ*), (2) the number of additional individuals infected per day by extra-network contacts during the ‘surge period’ (‘surge exposures’, *ε*), and (3) the maximum percent reduction in probability of one-time or casual sexual contact per day in response to perceived risk of mpox (‘behavioral adaptation’, ω). We fit these parameters to incident daily cases in NYC using a 5-step process.

First, we used Latin Hypercube Sampling (LHS) to generate 1,000 unique parameter sets, sampling between 40% and 100% transmission probability, between 100 and 1,000 surge exposures, and between 0% and 60% behavioral adaptation. Prior studies used transmission probability values below our minimum [12], but these lower values were based off estimates of household, rather than sexual, transmission, and do not generate sustained transmission in our network.

Second, we simulated a single run of outbreak dynamics under the baseline ‘first-dose priority’ strategy employed by NYC for each LHS-generated parameter set from May 14, 2022 to January 18, 2023 (250 simulated days). Third, we filtered out parameter sets which predicted a final cumulative case incidence outside of that observed in NYC +/-50%. Fourth, for each remaining parameter set, we calculated the likelihood that the simulated number of incident daily cases could generate the observed number of incident daily cases through Jan 18 using a negative binomial probability distribution. Finally, we drew 100 parameter sets (with replacement) with the calculated likelihood used as the relative draw probability for each parameter set. These 100 drawn parameter sets represent the posterior distributions of our fit parameters.

For each of these 100 parameter sets, we ran each of our vaccine model scenarios (Figure 1) from May 14, 2022 to May 14, 2023 and compared median and interquartile ranges of cumulative cases over time for each vaccine administration strategy. We additionally ran a third counterfactual model scenario where no vaccines were administered to measure the total percent of cases averted by vaccination compared to the ‘first-dose priority’ strategy (baseline model).

## Results

Our fitting procedure selected for a transmission probability of 59% [IQR: 55% – 65%], 440 surge exposures [IQR: 280 – 560], and behavioral adaptation of 37% [IQR: 26% – 47%] (Figure 2, Table 2). The posteriors of transmission probability and behavioral adaptation are both correlated with surge exposures (Figure S3). Thus, these parameter estimates should not be used in other contexts.

**Figure 2:**
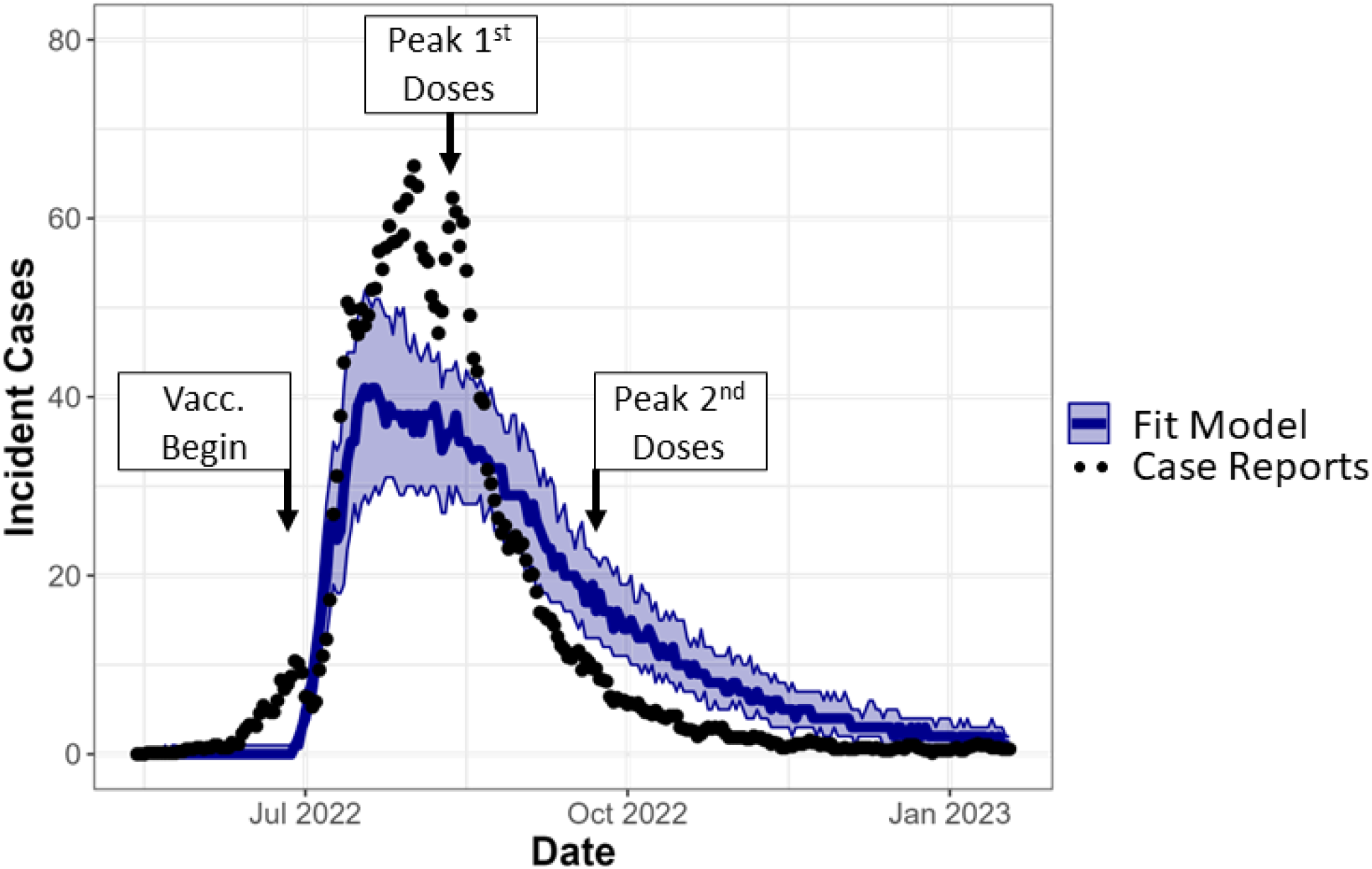
Mpox model fit to data. Dots indicate the 1 week running mean of incident daily cases in NYC, used as our fitting target. Blue line in represents median incident daily cases of model runs from the 100 parameter sets selected in our fitting procedure, with band representing interquartile range of model runs, assuming a ‘first-dose priority’ strategy. Labels indicate the beginning of pre-exposure vaccination on June 26^th^, peak administration of first doses, occurring the week of August 7^th^, and peak administration of second doses, occurring the week of September 18^th^.

**Table 1:**
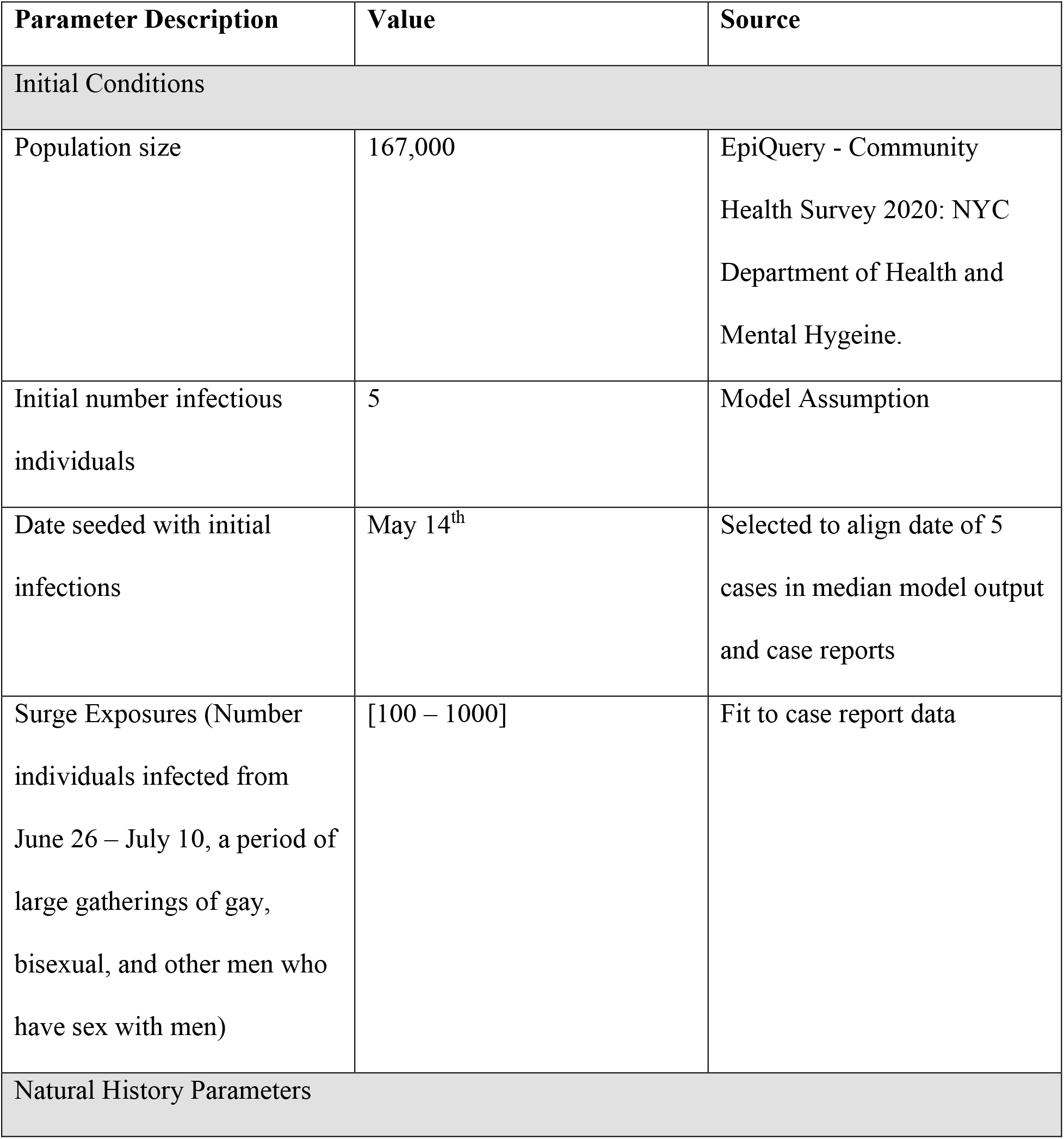
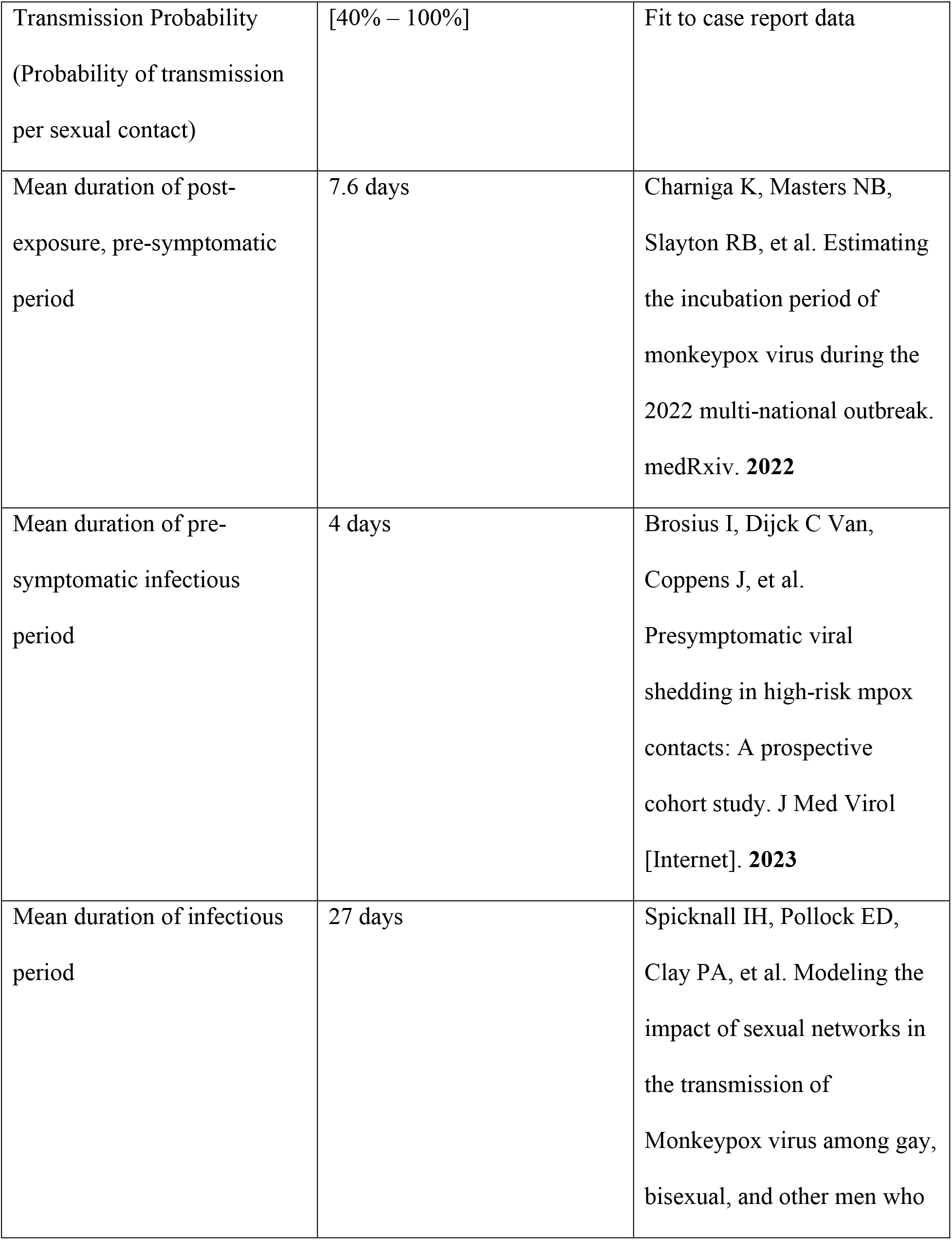
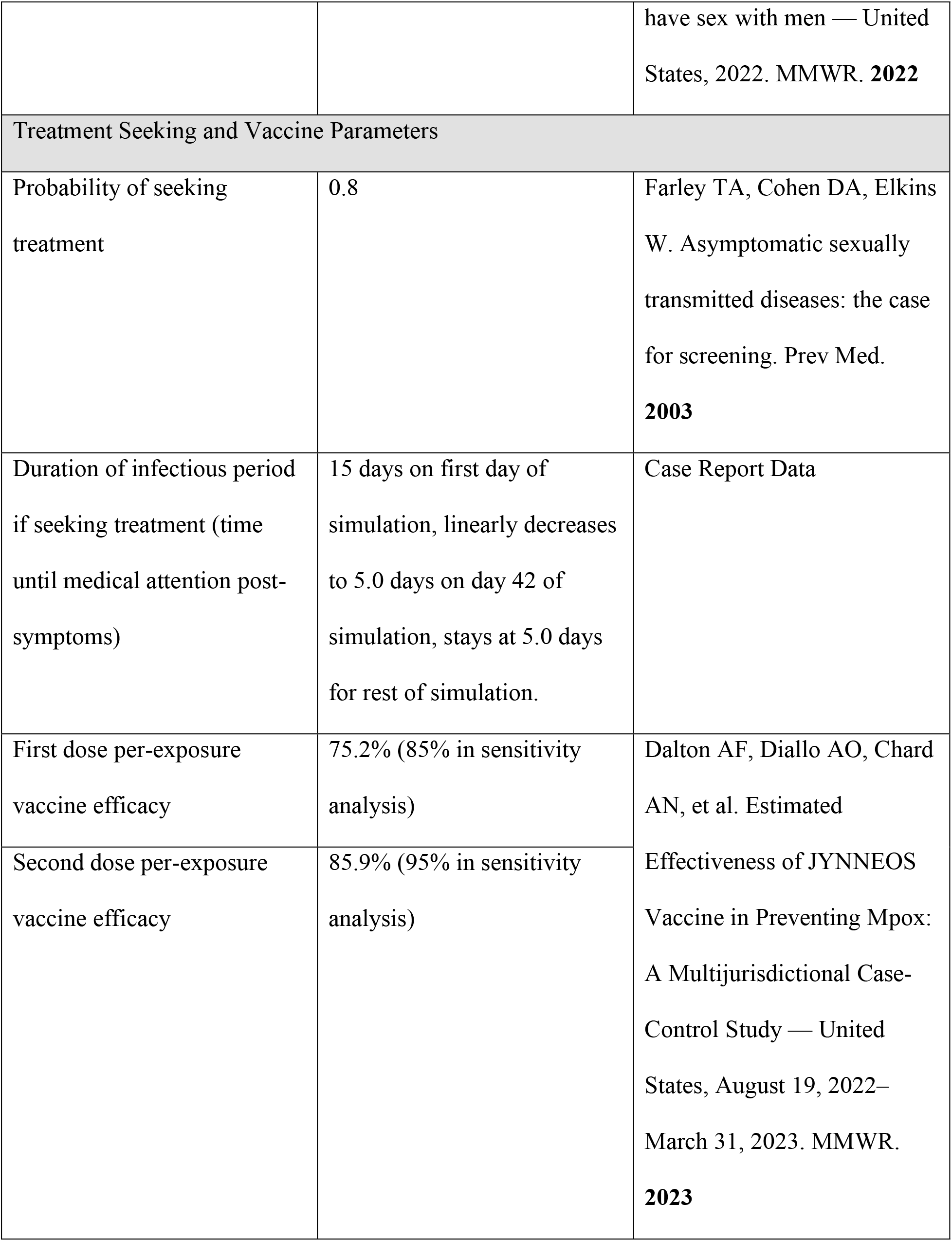
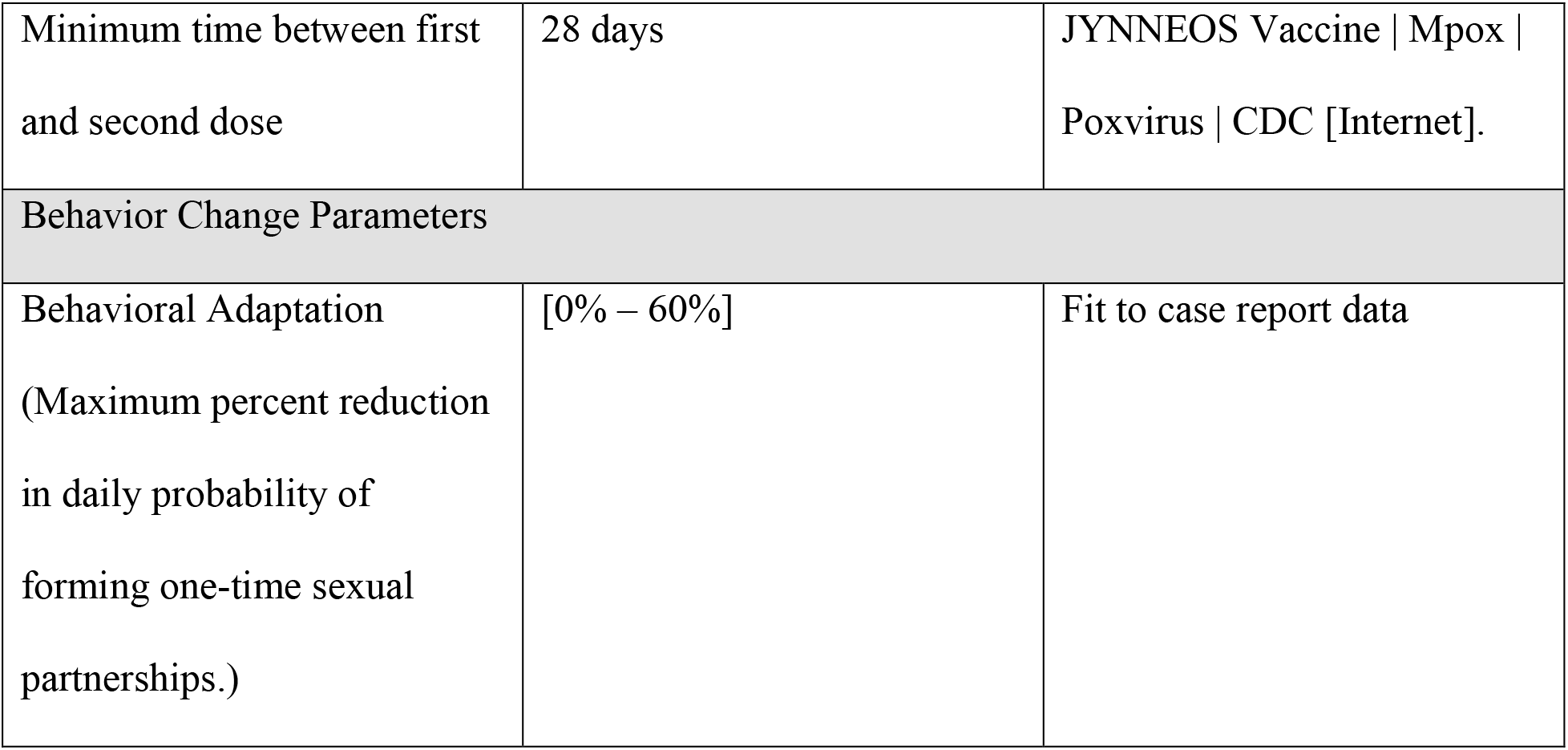
Model parameters. Values given as a range represent priors of parameters fit to data.

**Table 2:**
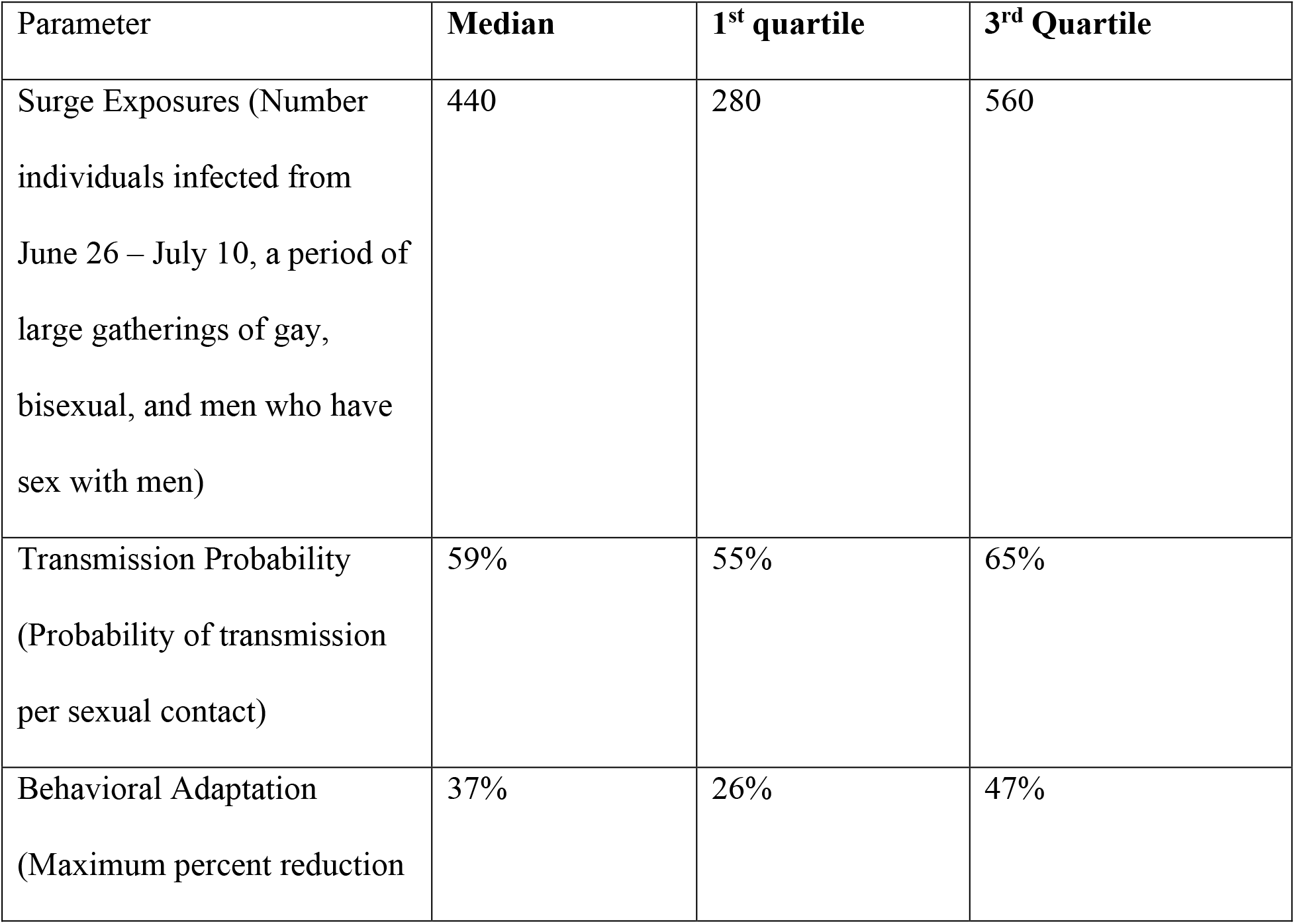
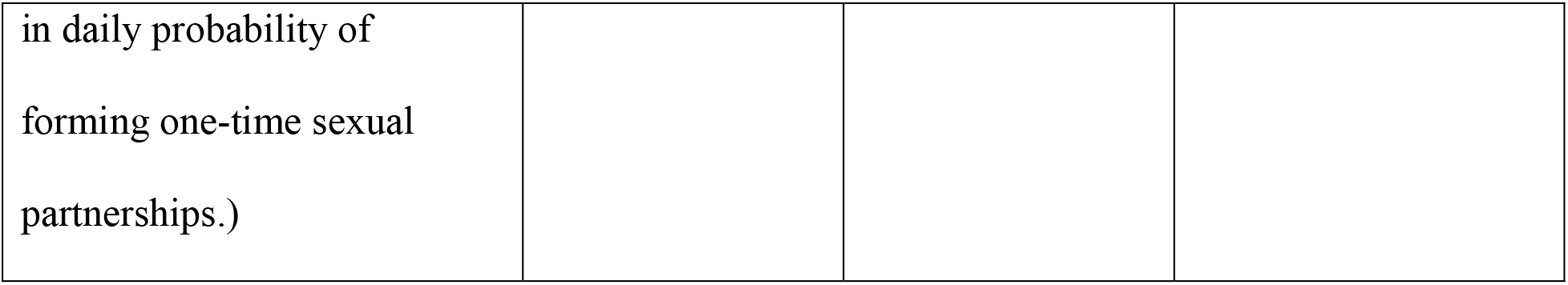
Posterior values of parameters fit to incident cases.

Our model estimates that there would have been 17,800 cumulative cases [IQR: 13,000 – 25,900] by May 14, 2023 in the counterfactual scenario with no vaccination. Our model further estimates that the baseline ‘first-dose priority’ strategy averted 81% [IQR: 75% – 86%] of these cases (Figure 3).

**Figure 3:**
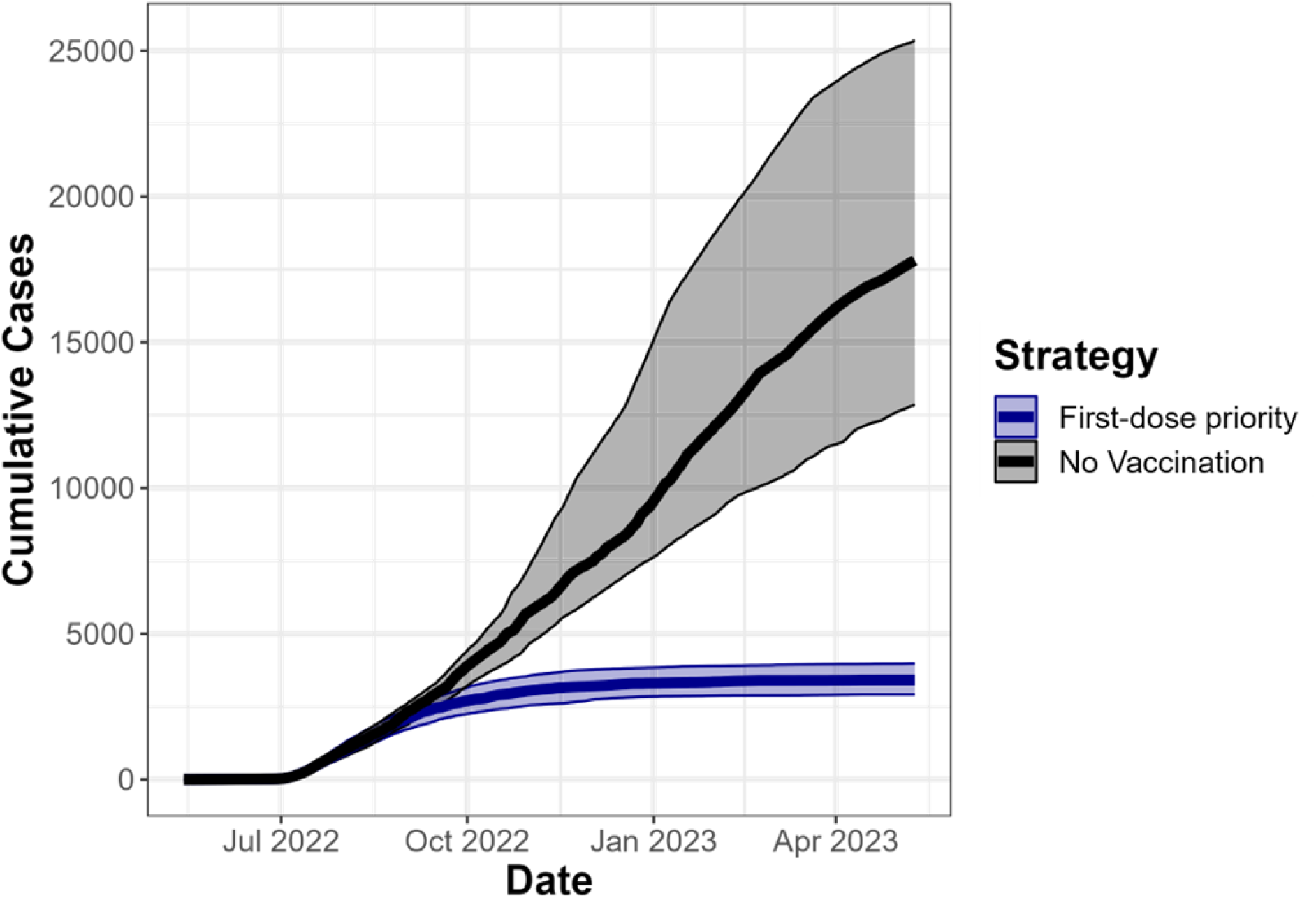
Estimated cumulative cases over a year under the ‘first-dose priority’ strategy employed by NYC compared to a ‘no vaccination’ scenario. Lines represent median cumulative cases of model runs from the 100 parameter sets selected in our fitting procedure, with bands representing interquartile ranges of model runs.

Taking the difference in cases averted across all parameter sets, we estimate that compared to the baseline ‘first-dose priority’ strategy employed by NYC, the counterfactual ‘intermediate’ strategy would have averted 3.0% [IQR: 1.2% – 4.5%] fewer cases, and the counterfactual ‘second-dose priority’ strategy would have averted 9.5% [IQR: 7.7% – 12%] fewer cases (Figure 4A). This would result in up to 2,000 fewer cases averted had NYC chosen an alternative vaccine administration strategy. This result is driven by exposure heterogeneity and incremental dose efficacy. In our model, the first individuals vaccinated have a one-time partner acquisition rate 250 times greater than individuals vaccinated later in the outbreak (Table S1), which favors narrow, 2 dose vaccination of individuals with higher sexual activity. However, this exposure heterogeneity is overwhelmed by the second vaccine dose adding relatively little incremental protection (10.7%) compared to the first dose (75.2%), which favors widespread, 1 dose vaccination.

**Figure 4:**
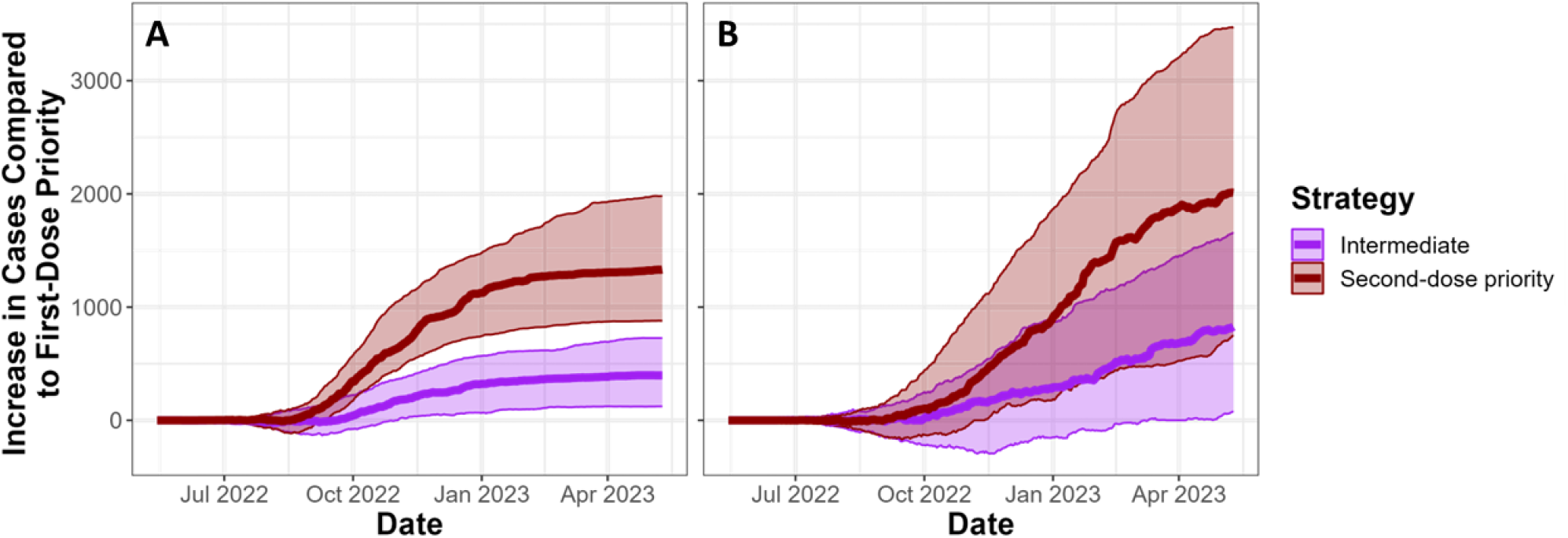
Comparison of different vaccine administration strategies*. We show the difference in cases under the ‘intermediate’ or ‘second-dose priority’ strategy compared to the ‘first-dose priority’ strategy on the Y-axis, over time on the X-axis. Solid lines indicate median values across fit parameter sets, and transparent bands indicate interquartile ranges across parameter sets. Panel A uses vaccine administration numbers shown in Figure 1, while panel B cuts doses given by 75% to emulate jurisdictions with lower dose availability. *First-dose priority: first and second doses in model are based on first and second doses administered by NYC. Intermediate: Individuals who are eligible for a second dose receive priority for available doses, but there is no preallocation. Second-dose priority: doses are preallocated to ensure that all individuals receive full course of thes vaccine.

NYC was allocated a relatively high number of JYNNEOS doses per high priority individual compared to many other jurisdictions. Thus, we conducted additional analyses, reducing doses administered in the NYC model by 75% to estimate whether the optimal vaccine administration strategy would differ if NYC had been allocated fewer doses. We found that in this scenario, the baseline ‘first-dose priority’ strategy was even more superior, averting 43% [IQR: 39% – 48%] of cases, while the counterfactual ‘intermediate’ strategy averted 11% [IQR: 2.0% – 18%] fewer cases, and the counterfactual ‘second-dose priority’ strategy averted 25% [IQR: 13% – 34%] fewer cases than the ‘first-dose priority’ strategy (Figure 4B).

In addition to incremental dose efficacy and heterogeneity in exposure probabilities, the overall transmissibility of a pathogen will influence whether widespread, 1 dose vaccination or narrow, 2 dose vaccination is favored. When we increase per-exposure vaccine efficacy (appendix S.5), our fitting procedure selected parameter sets with higher monkeypox virus transmissibility, increasing cumulative cases in the no-vaccine scenario by 8.2% (Figure 2 vs. Figure S5). Because most transmission events in our model happen among high-activity individuals, in this higher transmissibility scenario, high activity individuals may need more per-exposure protection to reduce the effective growth rate of mpox to below 1. This favors narrow, 2 dose vaccination strategies more than our primary analysis (Figure 2), resulting in all vaccine administration strategies averting similar numbers of cases (Figure S6).

## Discussion

During the 2022 mpox outbreak, demand for vaccines initially outstripped supply, preventing 2 dose vaccination for the total population likely to benefit from vaccination. Thus, NYC prioritized administration of first doses to reach a larger portion of the impacted population as quickly as possible. In this analysis, we modeled the strategy focusing on widespread, 1 dose vaccination employed by NYC, compared to counterfactual strategies focusing on 2 dose vaccination in narrower portions of the population. We estimated that the NYC vaccine rollout prevented 81% of potential mpox cases. We further estimated that NYC’s strategy of prioritizing widespread, 1 dose vaccination (the ‘first-dose priority’ strategy) averted more cases than alternative strategies prioritizing narrow, 2 dose vaccination, and would also have been the superior strategy had vaccine supply been even more limited.

Increasing vaccine coverage will decrease the probability of future mpox outbreaks and decrease the size of outbreaks if they occur [2]. CDC estimates that a vaccination threshold of 50% among priority populations (HIV PrEP indicated MSM and MSM living with HIV) would prevent monkeypox virus transmission [2]. However, 70% of analyzed jurisdictions, including urban centers with large MSM populations, are currently below this thresholds, with 34% of analyzed jurisdictions having vaccinated 25% or less of the priority population [2]. Thus, CDC recommends the continued vaccination of priority populations to prevent future mpox outbreaks [13]. Currently, vaccine availability outstrips demand among priority populations, and state and local health departments do not need to decide whether to prioritize narrow, 2 dose vaccination or widespread, 1 dose vaccination. Rather, state and local health departments can focus both on initiating the vaccine series for those in priority population who remain unvaccinated or have recently entered the priority population, followed by completion of the 2 dose vaccine series. Completing the vaccine series is important as the duration of effectiveness of a single dose is not known.

Given the potential for mpox resurgence as well as for mpox to spread to populations not previously impacted by mpox, demand for mpox vaccines could increase, potentially exceeding vaccine supply or vaccination capacity. In either situation, local and state health departments may consider whether to focus on prioritizing narrow, 2 dose vaccination, or widespread, 1 dose vaccination. Our analysis shows that in NYC, prioritizing widespread, 1 dose vaccination averted the greatest number of cases, making it the superior strategy, though it may be equivalent to other strategies if we are underestimating per-exposure vaccine efficacy. Even if all strategies were equivalent in averting cases, prioritizing widespread, 1 dose vaccination has the benefit of improving vaccine equity. In recent vaccine rollouts, groups who have historically faced systematic barriers to accessing healthcare received a disproportionally small number of initial vaccine doses [14]. Narrow, 2 dose vaccination strategies, which focus on concentrating all vaccine doses in a smaller number of individuals who access vaccine doses first, may thus exacerbate health inequities. This is particularly problematic as individuals are most likely to have close sexual contact with individuals of the same race or socioeconomic status [15]. Narrow, 2 dose strategies may thus lead to uncontrolled transmission in groups already facing a disproportionate burden of most health problems, exacerbating health disparities.

Vaccination can yield greater health benefits when the selection of vaccine administration strategies takes into account heterogeneity in immune function, which may in turn generate heterogeneity in both individual susceptibility and vaccine effectiveness. People with HIV (PWH) make up 19% of MSM [16,17], but 55% of mpox cases in the United States with known and recorded HIV status. The disproportionate representation of PWH among mpox cases may be caused by (a) PWH having a higher probability of mpox exposure, (b) PWH having a greater susceptibility to monkeypox virus infection, or (c) PWH being more likely to experience mpox symptoms when infected, given that coinfection between HIV and mpox can lead to severe, life-threatening outcomes in individuals not receiving antiretroviral therapy and who had low CD4 counts [18]. JYNNEOS® vaccination has reduced efficacy in individuals living with immunocompromising conditions, including uncontrolled HIV, with a reduction in first-dose effectiveness of 21.1%, and a reduction of second-dose effectiveness of 17.6%. Further, the incremental effectiveness of second dose vaccination is higher in people living with immunocompromising conditions than in others, with second dose vaccination increasing effectiveness by 20.8 percentage points, compared to 10.7 percentage points. Thus, because PWH may be more likely to acquire mpox, are more likely to experience severe symptoms, and benefit more from receiving a second vaccine dose than others, future vaccination strategies may benefit from giving PWH 2 dose vaccine courses as quickly as possible, even if the larger vaccine strategy is for widespread, 1 dose vaccination. This strategy could be paired with further efforts to connect people living with uncontrolled HIV to HIV services, given that increased CD4 counts may increase vaccine efficacy and reduce severity of mpox.

Choosing whether to prioritize completion of a multi-dose course in a narrow portion of the population at high likelihood of infection, versus widespread, 1 dose vaccination will arise in future outbreaks of other diseases, whenever vaccination involves multiple doses [19]. When choosing between vaccination strategies during an outbreak, public health agencies need to be quick, flexible, and make decisions with limited data. Mathematical modelers can conduct studies now, before those outbreaks occur, that will allow public health officials to make informed decisions during the initial stages of an outbreak. The optimal vaccine strategy in an outbreak will depend on multiple factors, including the incremental effectiveness of first vs. second doses, the population contact network, vaccine administration capacity, and the epidemic growth rate. Modeling studies can explore how these parameters shift the optimal vaccination strategy, while connecting these parameters to pre-existing data (e.g., the population contact network), or data that can be easily collected in the early stages of an outbreak (e.g. epidemic growth rate). Results from these studies can inform decisions for public health officials, leading to quick, evidence-based public health leadership.

## Acknowledgments

The findings and conclusions in this report are those of the authors and do not necessarily represent the views of the Centers for Disease Control and Prevention or the National Cancer Institute. We would like to thank Dr. Jennifer Rosen for her support.

## Data Availability

Data and code available upon reasonable request to authors.

## Notes

### Competing Interest Statement

The authors have declared no competing interest.

### Funding Statement

This study did not receive any funding

### Summary of Updates

Corrected spelling for name of co-author.

