## appendix for "Modeling the impact of prioritizing first or second vaccine doses during the 2022 mpox outbreak"

### S.1: Network parameters

Parameters for dynamic network were taken from a nationally distributed survey of sexual behaviors among MSM from 2017-2019 [5]. We recognize that sexual networks among MSM may have been impacted by Covid-19, but extensive egocentric surveys of sexual behavior among MSM taken after Covid-19 restrictions were implemented and relaxed have not been published. Thus, the ARTnet survey represents the most up to date and thorough source of sexual behavior data among MSM. Where necessary, ARTnet data was supplemented by prior surveys of sexual behavior among MSM in Atlanta [23] (Table S1).

ARTnet reports the proportion of MSM who have between 0 and 2 'Main' partners and between 0 and 3 'Casual' partners. Individuals with 2 'Main' partners represent a tiny portion of the population (e.g. individuals with 2 'Main' partners and 3 'Casual' partners make up 0.14% of the population). Modeling the behavior of tiny fractions of the population can lead to instability when fitting dynamic network models, so we removed individuals with 2 main partners from the model for this analysis (Table S1).

We extracted probabilities per day of engaging in one-time sexual partnerships from supplemental Figure 1 from [5], by taking the average mean of one-time daily partnership probabilities across percentages of the population corresponding to our sexual activity groups.

Table S1: Network parameters.

| **Parameter Description** | **Value** | **Source** |
|---|---|---|

| Sexual Network Parameters | | |
|---|---|---|
| Proportion individuals with 0 main and 0 casual partners | 0.30 | Weiss KM, Goodreau SM, Morris M, et al. Egocentric sexual networks of men who have sex with men in the United States: Results from the ARTnet study. Epidemics. **2020** |
| Proportion individuals with 0 main and 1 casual partners | 0.15 | |
| Proportion individuals with 0 main and 2 casual partners | 0.067 | |
| Proportion individuals with 0 main and 3 casual partners | 0.070 | |
| Proportion individuals with 1 main and 0 casual partners | 0.30 | |
| Proportion individuals with 1 main and 1 casual partners | 0.056 | |
| Proportion individuals with 1 main and 2 casual partners | 0.028 | |
| Proportion individuals with 1 main and 3 casual partners | 0.027 | |
| Mean duration of Main partnerships | 1,900 days | |
| Mean duration of Casual partnerships | 930 days | |
| Probability per day of individual in activity group 1 | 0 | |

| | |
|---|---|
| having a one-time sexual contact | |
| Probability per day of individual in activity group 2 having a one-time sexual contact | 0.0012 |
| Probability per day of individual in activity group 3 having a one-time sexual contact | 0.0052 |
| Probability per day of individual in activity group 4 having a one-time sexual contact | 0.016 |
| Probability per day of individual in activity group 5 having a one-time sexual contact | 0.064 |
| Probability per day of individual in activity group 6 having a one-time sexual contact | 0.30 |

| | | |
|---|---|---|
| Proportion of the population in activity groups 1-5 each | 0.19 | |
| Proportion of the population in activity group 6 | 0.05 | |
| Mean difference between square roots of ages of Main partners | 0.464 | Jenness SM, Weiss KM, Goodreau SM, et al. Incidence of Gonorrhea and Chlamydia Following Human Immunodeficiency Virus Preexposure Prophylaxis Among Men Who Have Sex With Men: A Modeling Study. Clin Infect Dis. **2017** |
| Mean difference between square roots of ages of Main partners | 0.586 | |
| Mean difference between square roots of ages of Main partners | 0.544 | |
| Proportion of population who are exclusively insertive | 0.242 | |
| Proportion of population who are exclusively receptive | 0.321 | |
| Proportion of population who are versatile | 0.437 | |
| Probability of having sexual contact per timestep with Main partner | 0.22 | |

| | |
|---|---|
| Probability of having sexual contact per timestep with Casual partner | 0.14 |
| Probability of having sexual contact per timestep with one-time partner | 1 |

**S.2: Calculating time between first and second dose administration when doses are pre-allocated**

Individuals receiving a full course of the JYNNEOS® vaccine are currently recommended to wait 4 weeks between doses. However, logistical issues often prevent patients from completing the two doses exactly 4 weeks apart, even when doses are available. Thus, for our ‘second-dose priority’ scenario, where doses were pre-allocated for individuals, we estimated the mean time that would pass between first and second doses in a pre-allocation scenario.

While NYC employed a ‘first-dose priority’ strategy, CDC guidance at the beginning of the mpox outbreak recommended that all individuals complete a full course of JYNNEOS at the recommended schedule. Thus, it is reasonable to assume that many jurisdictions pre-allocated a full 2-dose course to individuals. To parameterize time between doses in our ‘second-dose priority’, we measured the average time between doses in California, Washington D.C., and Illinois: three jurisdictions with large early mpox outbreaks.

We measured time between doses by fitting the transformed number of cumulative first doses administered over time to the number of cumulative second doses over time in each of these jurisdictions. We first multiplied the cumulative number of first doses administered over time by the percent of individuals receiving a first dose who eventually received a second dose in each jurisdiction. We treat the resulting data as the cumulative number of first doses administered to individuals who would eventually receive their second dose. We then shifted the cumulative number of first doses administered forward in time x = [1, 2… 10] weeks.  We calculate the sum of square differences between the first and second dose curves after shifting the first dose forward by x = [1, 2… 10]. We treat the value of x that results in the smallest sum of square differences as a proxy for mean time between first and second vaccine doses (Figure S1).

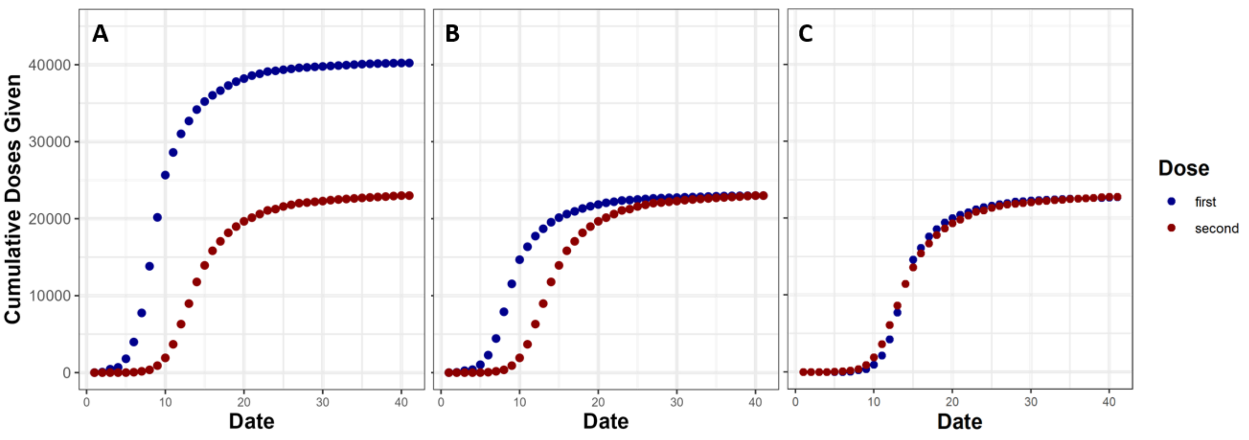

**Figure S1:** Visualization of method for estimating average time between first and second dose recipients.  In panel A, we show simulated cumulative first and second dose administration. In panel B, we multiply cumulative first doses by the proportion of individuals who go on to get second doses. In panel C, we shift the cumulative first dose curve forward by 5 weeks, minimizing sum of squared differences between the curves.

We estimate that on average 5 weeks passed between first and second vaccine dose administration for California, Illinois, and Washington D.C. We thus delay second dose vaccination by 5 weeks in our ‘second-dose priority’ scenario.

We note that our ‘second-dose priority’ scenario will overestimate the potential benefit of pre-allocating doses, because we do not account for individuals being pre-allocated doses and then being unable or unwilling to show up for their second dose.

**S.3: Calculating per-exposure vaccine efficacy**

Here, we calculate the per-exposure reduction in transmission probability due to vaccination, or per-exposure vaccine efficacy. The vaccine effectiveness (the percent reduction in infected individuals in the vaccinated vs. control arm of the trial, $E$), can be calculated as

$$E = 1 - \frac{1 - (1 - \mu(1 - \delta))^c}{1 - (1 - \mu)^c}$$

Via the probability of transmission per mpox exposure ($\mu$), the underlying per-exposure vaccine efficacy ($\delta$), and the number of infected individuals that people in an effectiveness trial contacted during the trial, filtering out those who had no exposures ($c$).

Thus, given a reported vaccine effectiveness, the per-exposure vaccine efficacy is given by

$$\delta = 1 - \frac{1 - (1 - (1 - E)(1 - (1 - \mu)^c))^{\frac{1}{c}}}{\mu}$$

If $c = 1$, then $\delta = E$. However, if $c > 1$ then $\delta > E$ (Figure S2).

For the purposes of this study, we assume that individuals in the JYNNEOS® effectiveness study [11] were exposed to mpox no more than once. However, it is likely that some individuals in the effectiveness study were exposed multiple times, and thus we underestimate per-exposure vaccine efficacy. Thus, we reran our analysis using higher vaccine efficacy estimates (S.5).

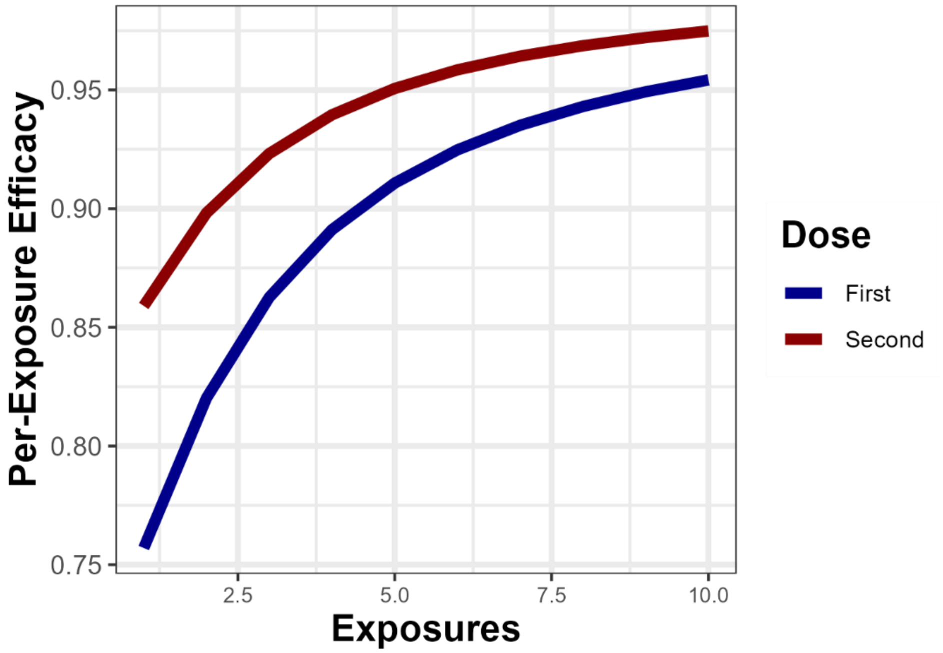

**Figure S2:** Here we show the derived per-exposure reductions in transmission probability due to one or two vaccine doses on the Y-axis, vs the number of assumed mpox exposures for vaccine effectiveness trial participants on the x-axis.

**S.4: Posterior distributions of fit model parameters**

Our fitting procedure drew 100 posterior parameter sets from our 1000 prior parameter sets, with the likelihood of a set being drawn being based on the negative binomial likelihood of the corresponding model output generating observed daily incident cases. We sampled with replacement, and thus our posterior distribution consists of 75 unique parameter sets.

In our prior parameter sets, all parameters were drawn from a uniform distribution, and parameters were not correlated with one another across sets. In our posterior distribution, we find that across parameter sets, per-exposure transmission probabilities are significantly negatively correlated with the number of individuals infected via extra-network contacts during the ‘surge period’ (‘surge exposures’) (Figure S3). We also find that maximum behavioral adaptation is positively correlated with ‘surge exposures’. This indicates that parameter sets with disparate parameter combinations can generate similar epidemic curves. E.g., a parameter set with low per-exposure transmission probability and a high number of ‘surge exposures’ could generate a similar epidemic curve as a parameter set with a high per-exposure transmission probability and a low number of ‘surge exposures’. This means that this analysis should not be used to infer the value of any one of these parameters individually.

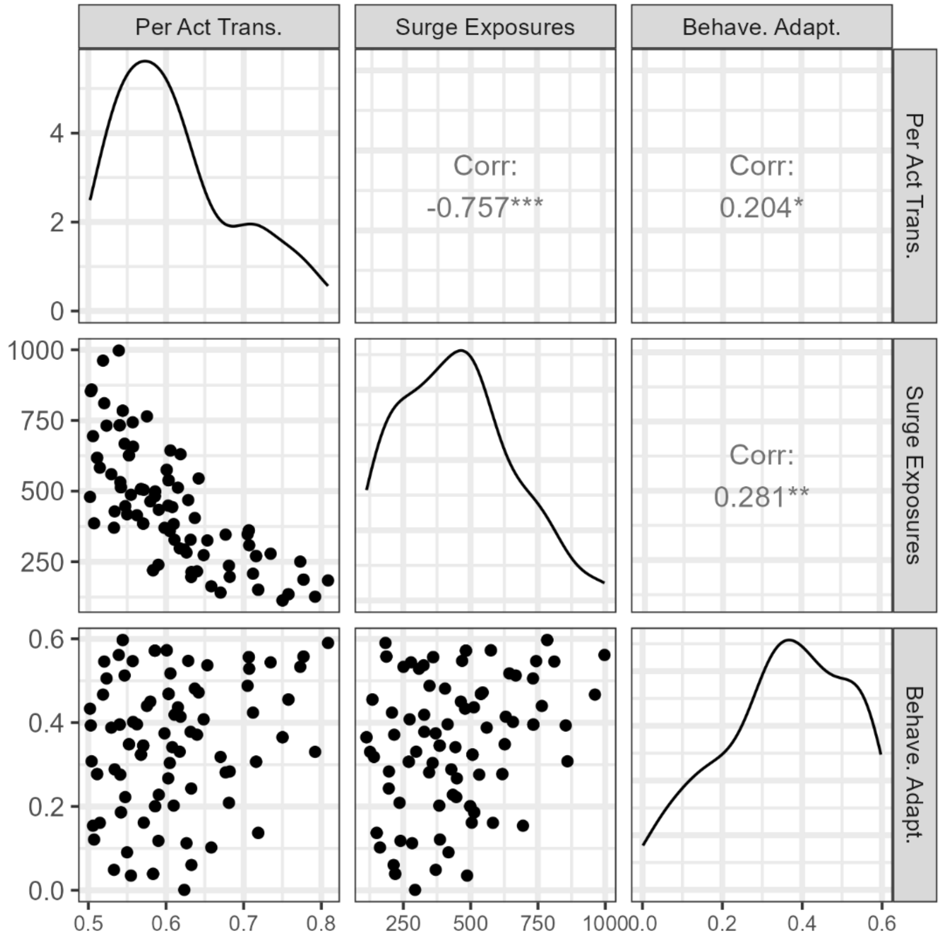

**Figure S3:** Here we show posterior distributions of the parameters in our model that were fit to incident case data, and correlations between those parameters. 'Per Act Trans.' indicates the probability of mokeypox virus transmission per contact between a susceptible and an infectious individual. 'Surge Exposures' indicates the number of individuals infected via extra-network contacts during the 'surge period' from June 26 - July 10. 'Behave. Adapt.' indicates behavioral adaptation, or the maximum percent reduction in one-time and casual sexual contacts due to the perceived risk of mpox. Panels show scatterplots of parameters across posterior parameter sets (bottom left panels), the posterior distribution of fit parameters (top-left to bottom-right), and correlations between parameters across posterior parameter sets (top-right panels).

**S.5: Sensitivity analysis of higher per-exposure vaccine efficacy**

Here, we redo analysis with higher per-exposure vaccine efficacy estimates (85% for first dose and 95% for second dose) to reflect uncertainty in translating vaccine effectiveness to per-exposure vaccine efficacy. We find that when running our analysis with these higher per-exposure vaccine efficacy values, The strategies do not dependably differ from one another in terms of cases averted (Figure S6).

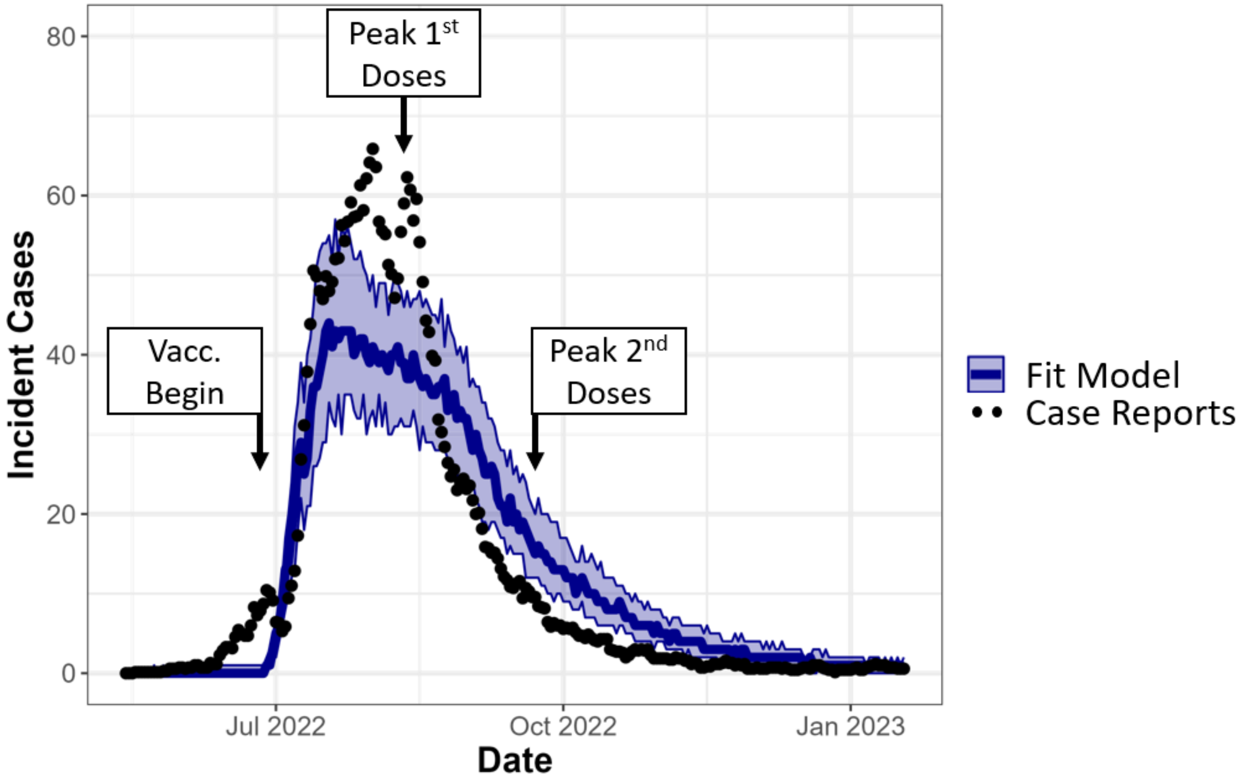

**Figure S4:** Repeat of Figure 2 with increased vaccine efficacy estimates. Mpox model fit to data. Dots indicate the 1 week running mean of incident daily cases in NYC, used as our fitting target. Blue line represents median incident daily cases of model runs from the 100 parameter sets selected in our fitting procedure, with band representing interquartile range of model runs, assuming a ‘first-dose priority’ strategy. Labels indicate the beginning of pre-exposure vaccination on June $26^{th}$, peak administration of first doses, occurring the week of August $7^{th}$, and peak administration of second doses, occurring the week of September $18^{th}$.

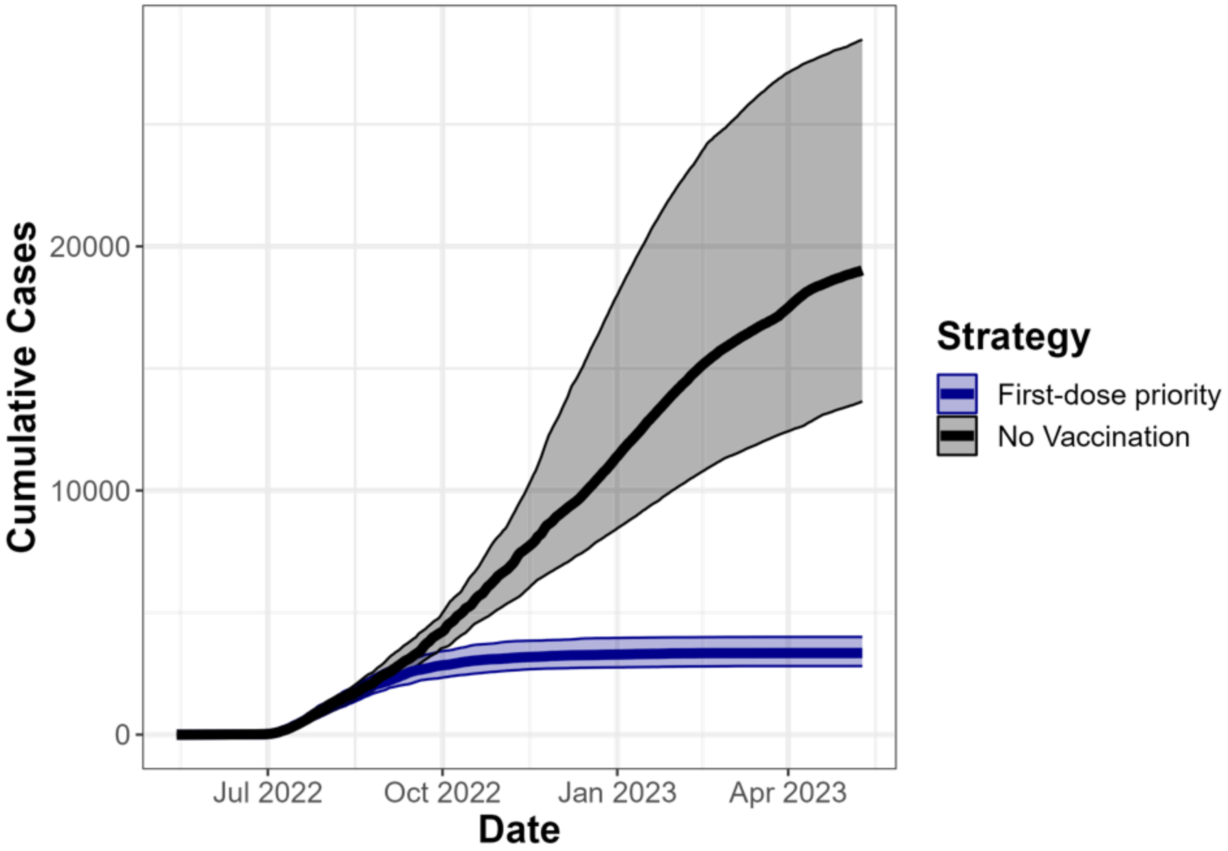

**Figure S5:** Repeat of Figure 3 with increased vaccine efficacy estimates. Estimated cumulative cases over a year under the ‘first-dose priority’ strategy employed by NYC compared to a ‘no vaccination’ scenario. Lines represent median cumulative cases of model runs from the 100 parameter sets selected in our fitting procedure, with bands representing interquartile ranges of model runs.

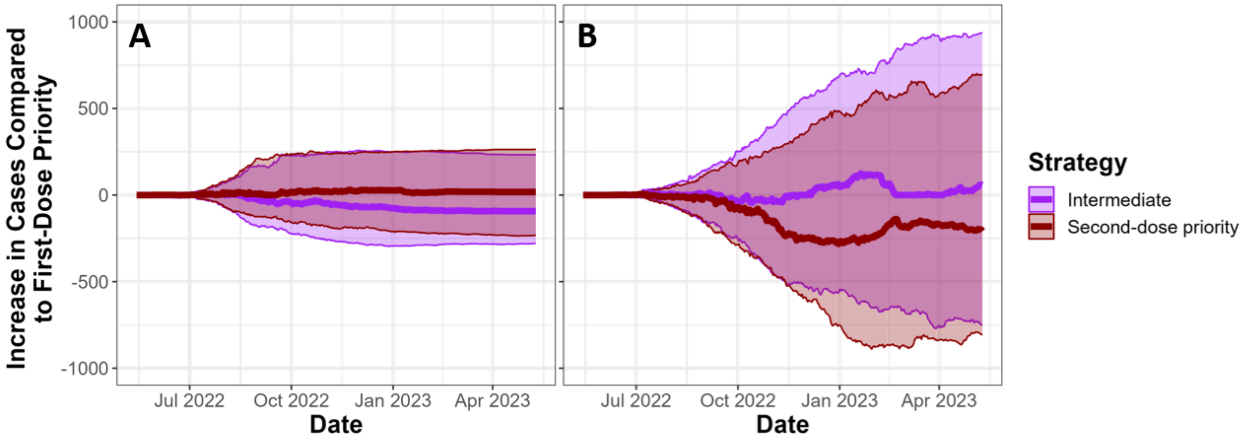

**Figure S6:** Repeat of Figure 4 with increased vaccine efficacy estimates. Comparison of different vaccine administration strategies[*]. We show the difference in cases under the ‘intermediate’ or ‘second-dose priority’ strategy compared to the ‘first-dose priority’ strategy on the Y-axis, over time on the X-axis. Solid lines indicate median values across fit parameter sets, and transparent bands indicate interquartile ranges across parameter sets. Panel A uses vaccine administration numbers shown in Figure 1, while panel B cuts doses given by 75% to emulate jurisdictions with lower dose availability. All interquartile values overlap 0 at the end of this timeseries, indicating that strategies do not dependably differ from one another in terms of cases averted.

[*]First-dose priority: first and second doses in model are based on first and second doses administered by NYC. Intermediate: Individuals who are eligible for a second dose receive priority for available doses, but there is no preallocation. Second-dose priority: doses are preallocated to ensure that all individuals receive full course of thes vaccine.
